# Practical Management of Adverse Events in Patients Receiving Tarlatamab, a DLL3-targeted Bispecific T-cell Engager Immunotherapy, for Previously Treated Small Cell Lung Cancer

**DOI:** 10.1101/2024.10.11.24315056

**Authors:** Jacob M. Sands, Stéphane Champiat, Horst-Dieter Hummel, Kelly G. Paulson, Hossein Borghaei, Jean Bustamante Alvarez, David P. Carbone, Jennifer W. Carlisle, Noura J. Choudhury, Jeffrey M. Clarke, Shirish M. Gadgeel, Hiroki Izumi, Alejandro Navarro, Sally C. M. Lau, Philip E. Lammers, Shuang Huang, Ali Hamidi, Sujoy Mukherjee, Taofeek K. Owonikoko

## Abstract

Tarlatamab is a bispecific T-cell engager (BiTE^®^) immunotherapy targeting delta-like ligand 3 (DLL3) and the cluster of differentiation 3 (CD3) molecule. In the phase 2 DeLLphi-301 trial of tarlatamab for patients with previously treated small cell lung cancer (SCLC), tarlatamab 10 mg every 2 weeks achieved durable responses and encouraging survival outcomes. Analyses of updated safety data from the DeLLphi-301 trial showed that the most common treatment-emergent adverse events (TEAEs) were cytokine release syndrome (CRS; 53%), pyrexia (38%), decreased appetite (36%), dysgeusia (32%), and anemia (30%). CRS was mostly grade 1 or 2 in severity, occurred primarily after the first or second tarlatamab dose, and was managed with supportive care, which included administration of antipyretics (e.g., acetaminophen), intravenous hydration and/or glucocorticoids. Other TEAEs of interest included neutropenia (16%) and immune-effector cell-associated neurotoxicity syndrome and associated neurologic events (10%). Given that tarlatamab is the first T-cell engager approved for the treatment of SCLC, raising awareness regarding the monitoring and management of tarlatamab-associated adverse events is essential. Here, we describe the timing, occurrence, and duration of these AEs and review the management and risk mitigation strategies used by clinical investigators during the DeLLphi-301 trial.

## Introduction

Tarlatamab is a bispecific T-cell engager (BiTE^®^) immunotherapy that binds both delta-like ligand 3 (DLL3) on cancer cells and the cluster of differentiation 3 (CD3) molecule on T cells. Tarlatamab causes T cell activation, release of inflammatory cytokines, and lysis of DLL3-expressing cells.^1^ Tarlatamab received accelerated approval from the US Food and Drug Administration in May 2024 for the treatment of patients with extensive-stage small cell lung cancer (ES-SCLC) with disease progression on or after platinum-based chemotherapy based on the durable responses and manageable safety profile seen in the DeLLphi-301 phase 2 trial.^2, 3^ Here, we provide an overview of the safety profile of tarlatamab 10 mg with updated data from the DeLLphi-301 trial (data cutoff: October 2, 2023). Details on the timing, occurrence, and duration of adverse events (AEs) are reported. We also provide clinical insights and strategies for AE management to optimize long-term patient treatment experience.

### Overview of the safety profile of tarlatamab in the DeLLphi-301 trial

In the phase 2 DeLLphi-301 trial in patients with previously treated SCLC, 133 patients received at least one dose of tarlatamab 10 mg. The most common treatment-emergent AEs (TEAEs; incidence ≥ 20% of patients) were cytokine release syndrome (CRS; 53%), pyrexia (38%), decreased appetite (36%), dysgeusia (32%), anemia (30%), constipation (29%), asthenia (25%), and fatigue (24%) (**Supplementary Table 1**). Other TEAEs of interest were neutropenia (16%) and immune effector cell-associated neurotoxicity syndrome (ICANS; 10%). The most common grade ≥ 3 TEAEs were lymphopenia (13.5%), anemia (8%), hyponatremia (6%), and neutropenia (6%) **(Supplementary Table 2)**. The time to onset of key AEs from the first tarlatamab dose is shown in **Figure 1**. CRS, fatigue, and ICANS were observed early, while neutropenia had a relatively delayed onset. **(Figure 1).** Treatment-related AEs led to dose interruption in 15% and discontinuation in 3% of patients.

**Figure 1.**
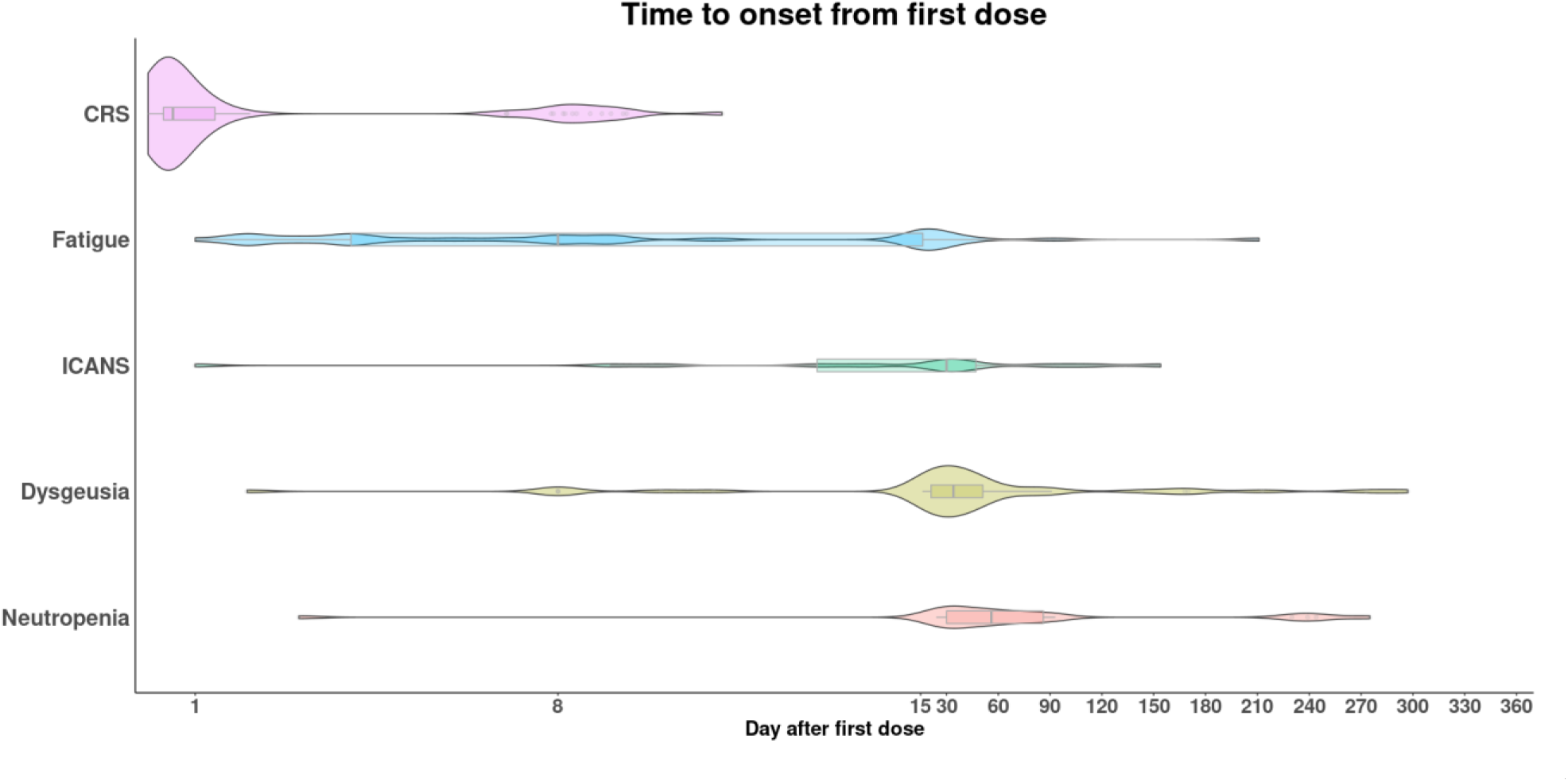
Time of AE onset from first dose. Violin-box plot showing the time to onset of some of the most common treatment-emergent adverse events from the first dose of tarlatamab in the DeLLphi-301 trial. The median time to onset and the interquartile range are shown within each plot, with the breadth of the violin-box plot indicating the patient distribution at any particular time point, and the height proportional to the incidence rate of each event. The x-axis has a variable scale. CRS: cytokine release syndrome; ICANS: immune effector cell-associated neurotoxicity syndrome and associated neurological events.

### Cytokine release syndrome (CRS)

Given the mechanism of action of BiTE immunotherapies, the safety profile of tarlatamab includes CRS.^4, 5^ CRS is a systemic inflammatory response caused by the activation of T cells and the subsequent release of inflammatory cytokines, such as interferon gamma.^4^ CRS diagnosis has often been considered a diagnosis of exclusion. Fever is considered the defining symptom of CRS, which can evolve to present with hypoxia, hypotension, tachypnea, headache, tachycardia, and rash (**Supplementary Table 3**).^6^ CRS can progress to include potentially life-threatening complications and end organ damage. This presents a diagnostic challenge because the differential diagnosis of fever is broad and can be caused by other conditions, particularly in patients with cancer, including, but not limited to infection, hypersensitivity, hemophagocytic lymphohistiocytosis, and tumor lysis syndrome.^7-10^ CRS is graded using the American Society for Transplantation and Cellular Therapy (ASTCT) consensus grading criteria based on the presentation of fever and the severity of hypoxia and hypotension.^6^ It should be noted, however, that the incidence and severity of CRS are lower with BiTE therapies relative to chimeric antigen receptor (CAR)-T-cell therapies.^11-15^

#### CRS in the DeLLphi-301 trial

In the DeLLphi-301 trial, CRS occurred in 53% of patients (70/133) treated with tarlatamab 10 mg (**Table 1**). CRS predominantly occurred after the first or second tarlatamab dose. CRS was reported in 41% of patients (54/133) following the first dose and in 29% (39/133) following the second dose (**Figure 2**). Three out of 133 patients (2%) experienced CRS at cycle 2 or later; each of these 3 patients had missed one or more doses during cycle 1 and had resumed treatment in cycle 2. The most common CRS symptom, besides fever, was hypotension (14/70; 20%) (**Table 1**). CRS was predominantly grade 1 (60%; 42/70) or grade 2 (fever with hypoxia and/or hypotension not requiring vasopressors) (39%; 27/70); one patient (1.4%) experienced grade 3 CRS (**Table 2**). There were no grade 4 or 5 CRS events. The median time to CRS onset from the first tarlatamab dose was 13.7 hours (interquartile range [IQR], 9.3–33.2) (**Figure 1**). The median time to resolution (Kaplan-Meier estimate) was 3 days (IQR, 2–5). The median time to onset of grade ≥ 2 CRS from hours, regardless of whether it subsided on antipyretic therapy, were more likely to experience a grade 2 or higher CRS event.

**Figure 2:**
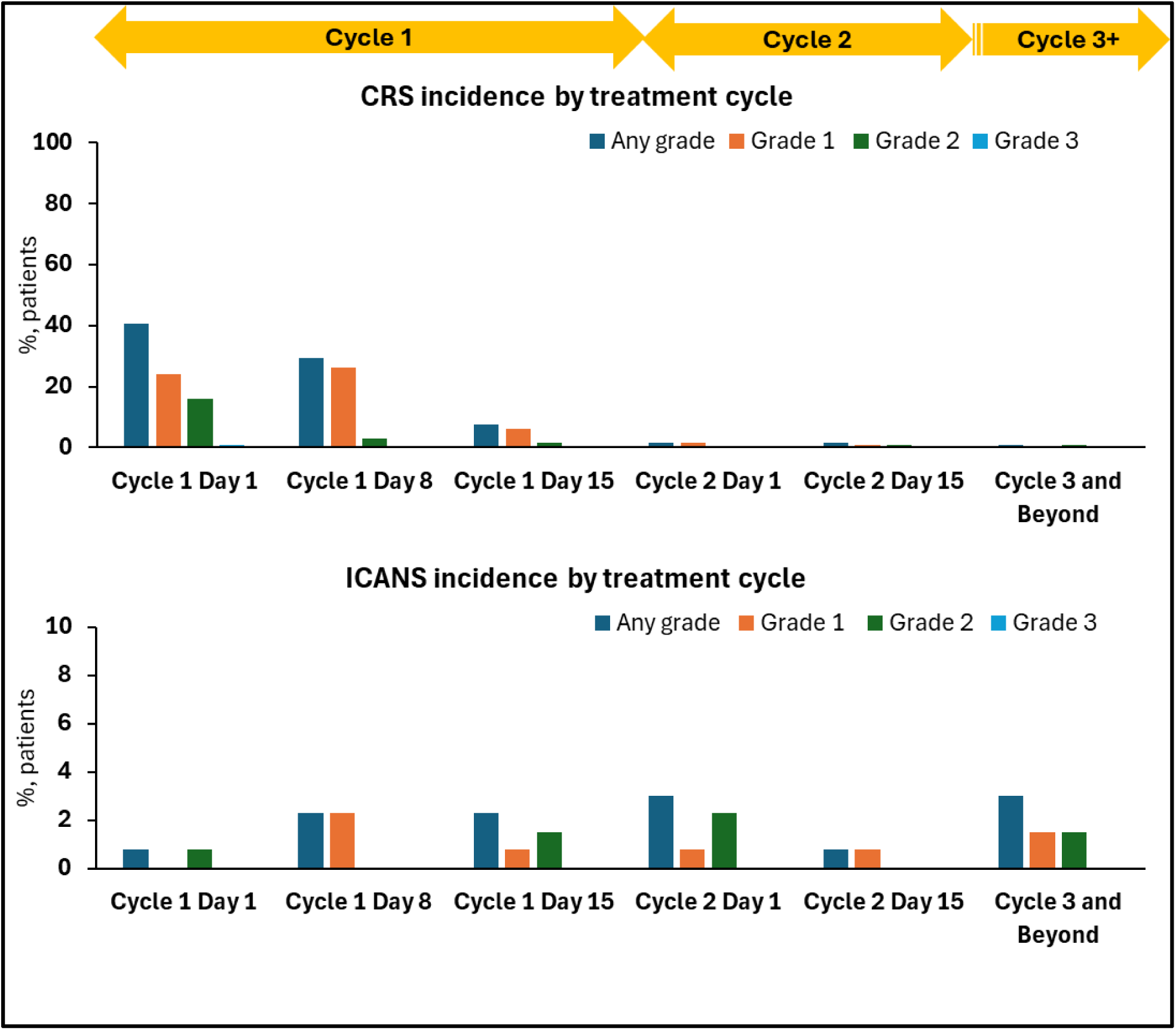
Incidence of CRS and ICANS by cycle and grade. **The incidence of CRS and ICANS according to severity and treatment cycle are shown for patients in the 10 mg tarlatamab dose cohort.** During the first cycle, CRS was reported in 41% of patients (54/133) following the first tarlatamab dose (days 1–7), in 29% (39/133) following the day 8 dose (days 8–14), and 8% (10/133) after the day 15 dose (days 15–28). CRS events were identified on the basis of a narrow search for preferred terms in MedDRA v 26.1 and were graded according to the American Society for Transplantation and Cellular Therapy 2019 consensus criteria. During the first cycle, ICANS was reported in <1% of patients (1/133) following the first tarlatamab dose (days 1–7), in 2% (3/133) following the day 8 dose (days 8–14), and in 2% (3/133) of patients after the day 15 dose (days 15–28). ICANS data include associated neurological events identified on the basis of a broad search for 61 preferred terms in MedDRA. CRS: cytokine release syndrome; ICANS: immune effector cell-associated neurotoxicity syndrome and associated neurological events; MedDRA: Medical Dictionary for Regulatory Activities

**Table 1.** Incidence of CRS and ICANS symptoms in the DeLLphi-301 trial.

| **CRS/ICANS symptoms, n (%)<sup>a</sup>** | **Tarlatamab 10 mg**<br>**(N = 133)** |
| --- | --- |
| ***Number of patients reporting CRS*** | ***70 (52.6)*** |
| Any CRS symptom | 70 (100.0) |
| Fever (temperature $\geq 38^{\circ}\text{C}$) | 68 (97.1) |
| Hypotension | 14 (20.0) |
| Hypoxia | 11 (15.7) |
| Headache | 9 (12.9) |
| Nausea | 7 (10.0) |
| Fatigue | 7 (10.0) |
| Tachycardia | 6 (8.6) |
| Rigors | 6 (8.6) |
| Myalgia | 5 (7.1) |
| Vomiting | 4 (5.7) |
| Malaise | 3 (4.3) |
| Arthralgia | 3 (4.3) |
| Anorexia | 2 (2.9) |
| Tremor | 2 (2.9) |
| ***Number of patients reporting ICANS<sup>b</sup>*** | ***7 (5.3)*** |
| Any ICANS symptom | 6 (85.7) |
| Confusion | 4 (57.1) |
| Impaired attention | 3 (42.9) |
| Motor weakness/findings | 1 (14.3) |
| Dysgraphia | 1 (14.3) |
| Increased tone | 1 (14.3) |

The safety analysis set includes all patients who received at least 1 dose of tarlatamab.
<sup>a</sup>Percentage for patients reporting CRS/ ICANS events are based on number of patients in the safety analysis set (N). Percentages for patients reporting symptoms are calculated using the number of patients reporting CRS or ICANS events as denominator.
<sup>b</sup>Patients with ICANS-only symptoms. Patients with associated neurological events were not included here.
ALT: alanine aminotransferase; AST: aspartate aminotransferase; CRS: cytokine release syndrome; ICANS: immune effector cell-associated neurotoxicity syndrome.

**Table 2.**
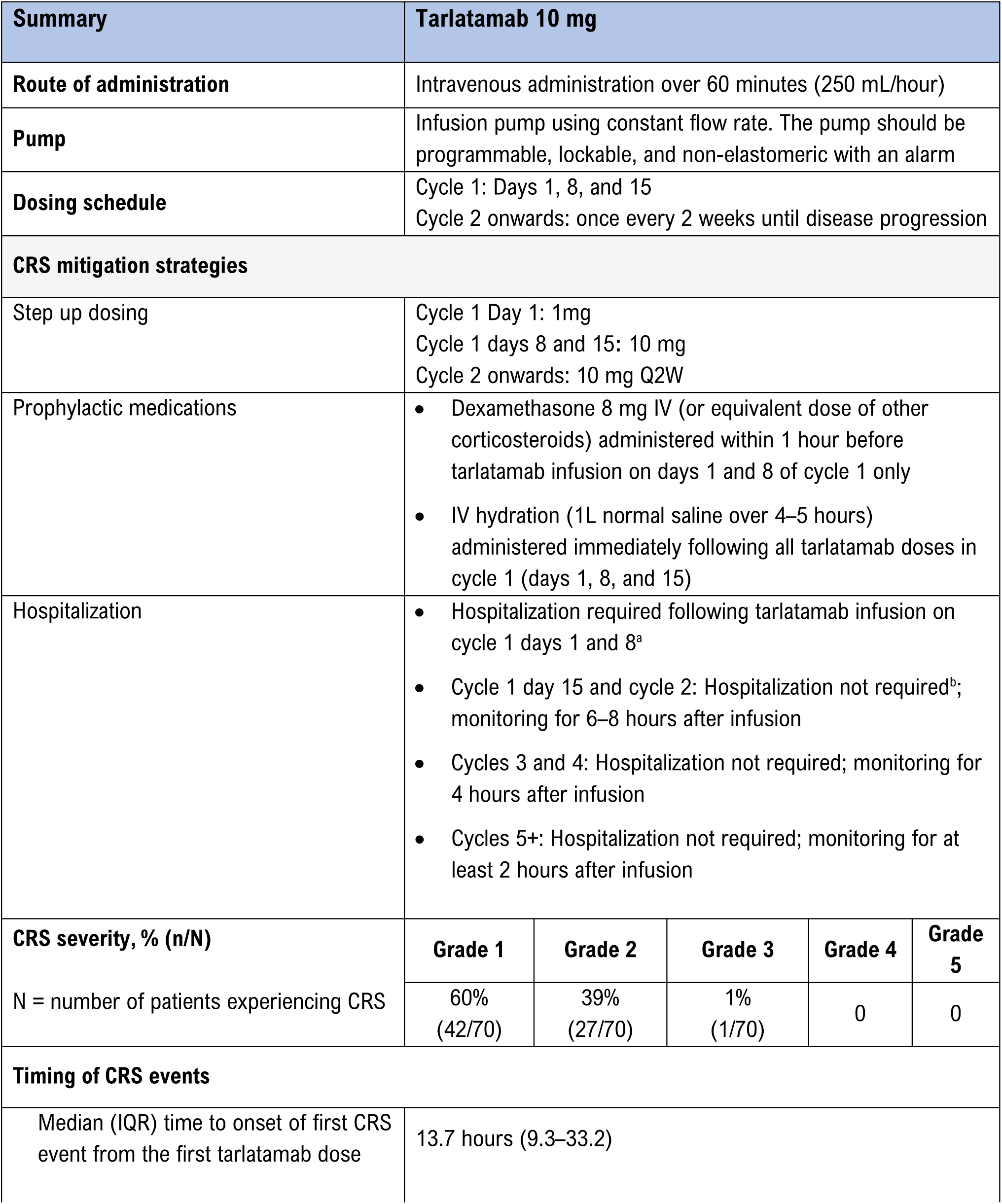
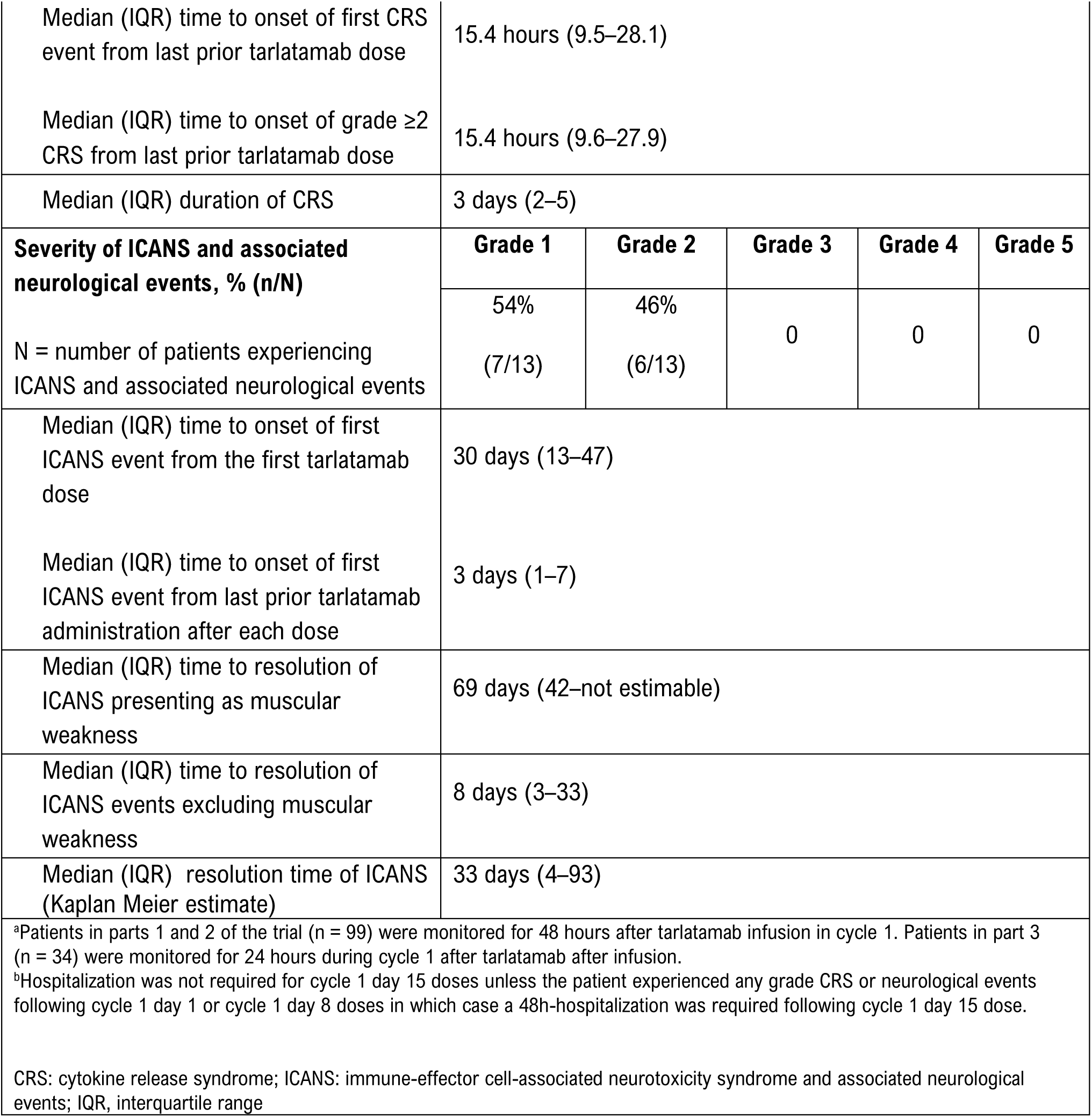
Summary of tarlatamab administration, CRS mitigation strategies, timing, and incidence of CRS and ICANS in the DeLLphi-301 trial.

Recurrent CRS occurred in 25% of patients (33/133). All patients with recurrent CRS had experienced their first CRS event during cycle 1. Recurrent CRS primarily occurred in cycle 1 and was grade 1 (88%) or grade 2 (12%) **(Table 3)**.

**Table 3.**
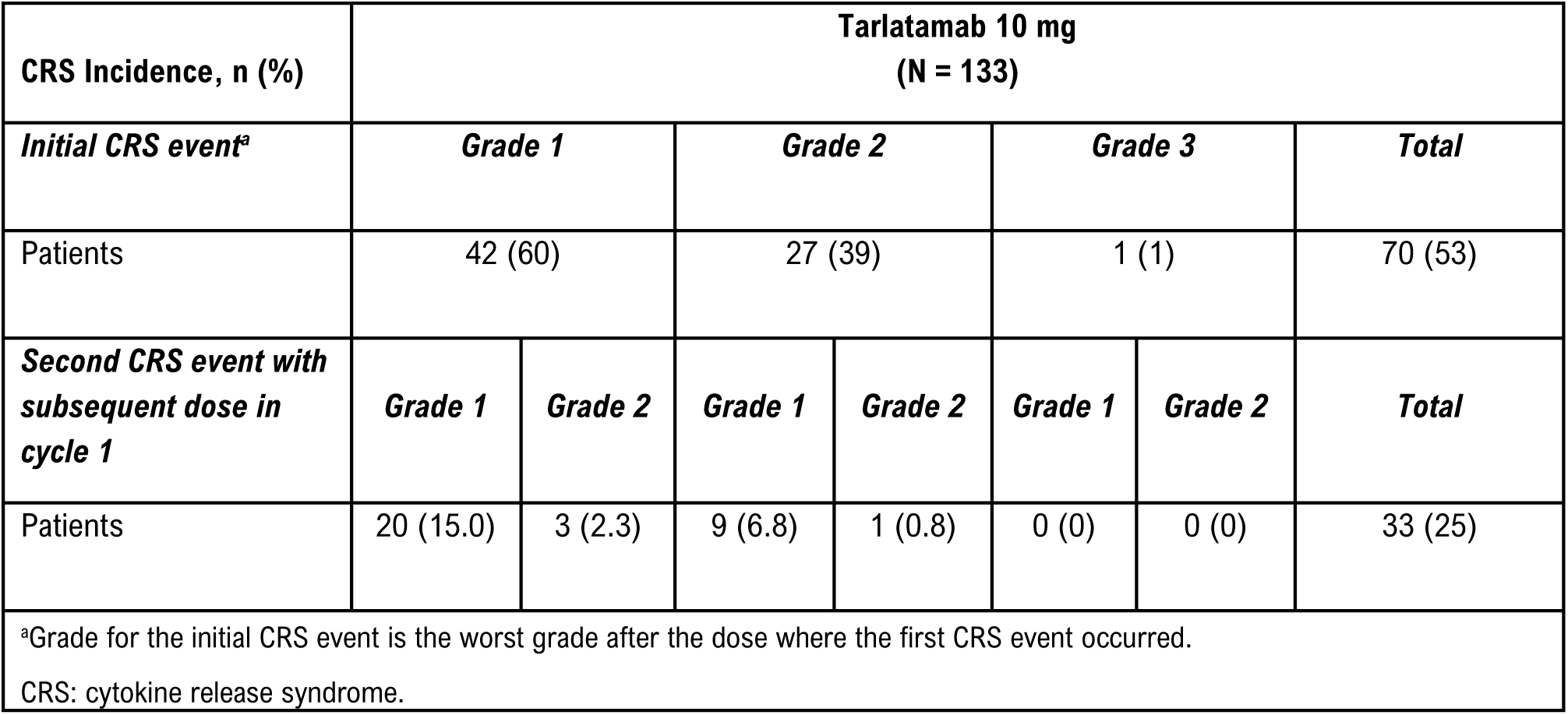
Frequency of patients experiencing > 1 CRS event in the DeLLphi-301 trial.

CRS led to dose interruption in 4 of 70 patients (6%). No patient discontinued tarlatamab due to CRS. One patient with concurrent grade 3 CRS died from respiratory failure. The respiratory failure was assessed by the investigator to be treatment-related, although this categorization was confounded by the presence of other contributing factors, including baseline chronic obstructive pulmonary disease requiring supplemental oxygen, baseline compromised pulmonary functional reserve, concurrent grade 3 CRS and pneumonitis after cycle 1 day 1 (C1D1) treatment, and a decision against escalation to intensive care unit (ICU) level of care.

#### CRS management in the DeLLphi-301 trial

When a patient presented with fever (≥ 38°C), a thorough work-up to rule out other causes (infection or sepsis) was performed. If CRS was suspected, the CRS event was graded according to the ASTCT Consensus Grading and treated based on the grade (**Table 4)**.^2, 6^ The management of CRS in the DeLLphi-301 trial protocol is described in detail in **Table 4**. Of note, clinical trial sites were required to have medications for treating CRS available on-site, including glucocorticoids and tocilizumab or siltuximab, if tocilizumab was not available.

**Table 4.** Management approach for CRS as used in the DeLLphi-301 trial protocol.

| CRS Grade<sup>a</sup> | Defining Symptoms | Management<sup>a</sup> | Dose Modification |
| --- | --- | --- | --- |
| **Grade 1** | Symptoms require symptomatic treatment only<br>Fever $\geq 38^{\circ}\text{C}$, without hypotension or hypoxia | Symptomatic treatment (e.g., paracetamol/acetaminophen) for fever | No treatment interruption needed |
| **Grade 2** | Symptoms require and respond to moderate intervention.<br>Fever $\geq 38^{\circ}\text{C}$,<br>Hypotension not requiring vasopressors, and/or<br>Hypoxia requiring low-flow nasal cannula or blow-by | Manage as for grade 1<br>Supplemental oxygen when oxygen saturation $< 90\%$ on room air<br>IV fluids when SBP $< 85$ mmHg and/or persistent tachycardia (heart rate $> 120$ bpm) or symptomatic (eg, dizziness/lightheadedness)<br>Consider a single dose of dexamethasone 8 mg IV (or equivalent)<br>Consider tocilizumab<sup>b</sup> or equivalent) | Interrupt tarlatamab until event improves to CRS grade $\leq 1$, then restart tarlatamab at the same dose level. Discontinue if there is no improvement to CRS ≤ grade 1 within 7 days |
| **Grade 3** | Severe symptoms defined as temperature $\geq 38^{\circ}\text{C}$ with:<br>Hemodynamic instability requiring a vasopressor (with or without vasopressin) or<br>Worsening hypoxia or respiratory distress requiring high-flow nasal canula ($> 6$ L/min oxygen) or face mask | Manage as for grade 2<br>Supplemental oxygen<br>Vasopressor ± vasopressin<br>IV dexamethasone (or equivalent), and/or<br>Tocilizumab<sup>b</sup> (siltuximab [11 mg/kg] administered over 1 hour as an intravenous infusion, may be used if tocilizumab is not available) | Interrupt tarlatamab for at least 3 days until the event improves to CRS grade $\leq 1$<br>If there is no improvement to grade $\leq 1$ within 7 days, or grade 3 toxicity re-occurs within 7 days of re-initiation, permanently discontinue tarlatamab |
| **Grade 4** | Life-threatening symptoms defined as temperature $\geq 38^{\circ}\text{C}$ with:<br>Hemodynamic instability requiring multiple vasopressors (excluding vasopressin)<br>Worsening hypoxia or respiratory distress despite oxygen administration requiring positive pressure (eg, CPAP, BiPAP, intubation, and mechanical ventilation) | Manage as for grade 3<br>Supplemental oxygen (positive pressure)<br>Multiple vasopressors<br>IV dexamethasone (or equivalent)<br>Tocilizumab<sup>b</sup> (siltuximab [11 mg/kg] administered over 1 hour as an intravenous infusion, may be used if tocilizumab is not available) | Permanently discontinue tarlatamab |

<sup>a</sup>Based on the ASTCT Consensus Grading (2019).<sup>6</sup>
<sup>b</sup>In the DeLLphi-301 trial, the recommended tocilizumab dose was 8 mg/kg IV over 1 h (not exceeding 800 mg/dose). Tocilizumab was repeated every 8 hours as needed if patient was not responsive to IV fluids or to increasing supplemental oxygen and limited to a maximum of three doses in a 24-hour period with a maximum total of four doses.

| **CRS**<br>**Grade<sup>a</sup>** | **Defining Symptoms** | **Management<sup>a</sup>** | **Dose Modification** |
| --- | --- | --- | --- |
| ASTCT, the American Society for Transplantation and Cellular Therapy; BiPAP: bilevel positive airway pressure; CPAP: continuous positive airway pressure; CRS, cytokine response syndrome; IV = intravenous; SBP: systolic blood pressure. |  |  |  |

As CRS in the DeLLphi-301 trial was predominantly grade 1 or grade 2 in severity, it was generally managed with supportive care that included antipyretics (e.g., acetaminophen), intravenous (IV) hydration, and/or glucocorticoid use. Additional interventions such as tocilizumab (used in 8/70 of patients), supplemental high-flow oxygen (> 6 L/min) (1/70 patients), and/or vasopressor support (1/70 patients) were infrequently required (**Supplementary Table 4**); siltuximab was not used. Incidence and duration of hospitalization for serious CRS

### Immune effector cell-associated neurotoxicity syndrome (ICANS)

The pathophysiology of ICANS remains poorly understood. Increased production of proinflammatory cytokines may lead to the activation of endothelial cells, increased permeability of the blood brain barrier, and elevated levels of cytokines in the cerebrospinal fluid.^16^ Signs and symptoms are variable, and may include aphasia, altered level of consciousness, impairment of cognitive skills, motor weakness, seizures, and cerebral edema.^6^ Per the ASTCT grading system, ICANS is graded using a combination of the immune effector cell-associated encephalopathy (ICE) score, level of consciousness, motor findings, seizure assessment, intracranial pressure level or extent of cerebral edema (**Supplementary Tables 5 and 6**).^6^ ICANS is also a diagnosis of exclusion. Symptoms of ICANS can appear similar to symptoms observed with infection/sepsis, stroke, paraneoplastic syndromes, use of pain medications, and new or progressive disease in the central nervous system (CNS).^7^

#### ICANS in the DeLLphi-301 trial

As ICANS can have a wide range of manifestations with non-specific symptoms, associated neurologic events were included in the reporting of ICANS^17^ (see **Supplementary Table 7** for the list of associated neurologic events). In the DeLLphi-301 trial, ICANS and associated neurologic events (henceforth referred to as ICANS) occurred in 10% of patients (13/133) treated with tarlatamab 10 mg (**Table 2**). Patients with ICANS reported symptoms which included confusion, impaired attention, motor weakness/findings, dysgraphia, and increased tone **(Table 1)**. All ICANS events were grade 1 (54% of patients; 7/13) or grade 2 (46%; 6/13) in severity (**Table 2**) and mostly occurred during cycle 1 or 2**. (Figure 2).** Of the 13 patients with ICANS, the median time to onset from the first tarlatamab dose was 30 days (IQR, 13–47) and the median time to resolution was 33 days (IQR, 4–93) (Kaplan-Meier estimate) (**Figure 1**). ICANS events that included confusion or impaired attention symptoms (11/16) were shorter in duration (median time to resolution of 8 days [IQR, 3–33]), whereas ICANS events characterized by low grade muscular weakness symptoms (5/16) generally took longer to resolve (median time to resolution: 69 days [IQR: 42–not estimable]).

ICANS led to dose interruption in one patient and discontinuation in one patient. No deaths due to ICANS were reported. Two patients were hospitalized for serious ICANS following the first or second dose of tarlatamab, and two patients were hospitalized for serious ICANS following the third or later dose of tarlatamab. The frequency and duration of hospitalization for serious ICANS are shown in **Table 5**. In a subgroup analysis, ICANS occurred in 3 of 22 patients (14%) with brain metastases and 5 of 77 patients (6%) without brain metastases.^18^ All events were grade 1 or 2 in severity.

**Table 5.**
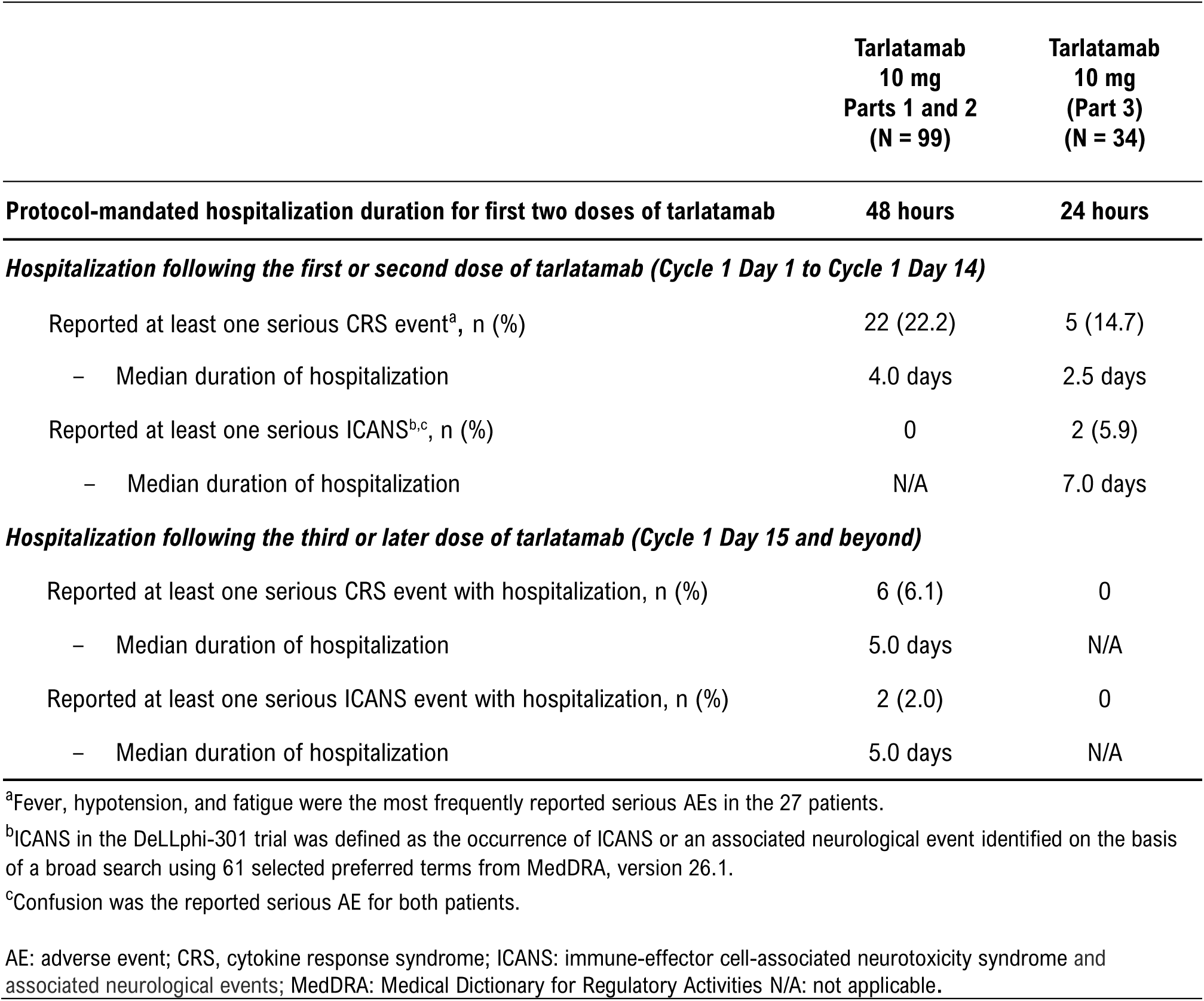
Hospitalization details for patients experiencing serious CRS and ICANS in the DeLLphi-301 trial.

#### ICANS management in the DeLLphi-301 trial

Patients in the DeLLphi-301 trial were monitored for clinical signs or symptoms that could be suggestive of ICANS including confusion, impaired attention, aphasia, tremor, and muscle weakness. It was important to rule out other neurological etiologies, such as epilepsy, infectious disease, progression of CNS metastasis, side effects of other concomitant medications, or electrolyte disturbances within or outside the context of paraneoplastic syndromes. In the DeLLphi-301 trial, neurological exams were performed as clinically indicated on or after dose administration. Although there were no grade 3 ICANS events in this trial, the protocol recommended a neurology consultation at the first sign of neurotoxicity and brain imaging for patients with persistent grade ≥ 3 neurotoxicity, with consideration for repeat neuroimaging (computed tomography [CT] or magnetic resonance imaging [MRI]) every 2–3 days. Electroencephalograms were required for the assessment of any seizure activity that was grade ≥ 2. If ICANS was suspected, the next step involved using the ICE assessment tool (**Supplementary Table 6**) and the ASTCT ICANS consensus grading criteria (**Supplementary Table 5**). Management strategies for ICANS used in the DeLLphi-301 trial are listed in **Table 6**. Briefly, grade 1 and grade 2 ICANS were to be managed by supportive care (grade 1) or steroids (grade 2). Steroids that were used included dexamethasone (10 mg IV), repeated every 6 hours if symptoms worsened, or methylprednisolone (1 mg/kg IV every 12 hours if symptoms worsened). For more severe events (grade ≥ 3) respiratory assistance with mechanical ventilation for airway protection (for grade 3 events), and treatment of status epilepticus per institutional guidelines (for grade 4 events) were recommended, though this was never required in the DeLLphi-301 trial (see **Table 6** for details). If grade ≥ 3 CRS with hypotension was also present, tocilizumab 8 mg/kg IV over 1 hour (not exceeding 800 mg/dose) was considered.

**Table 6.**
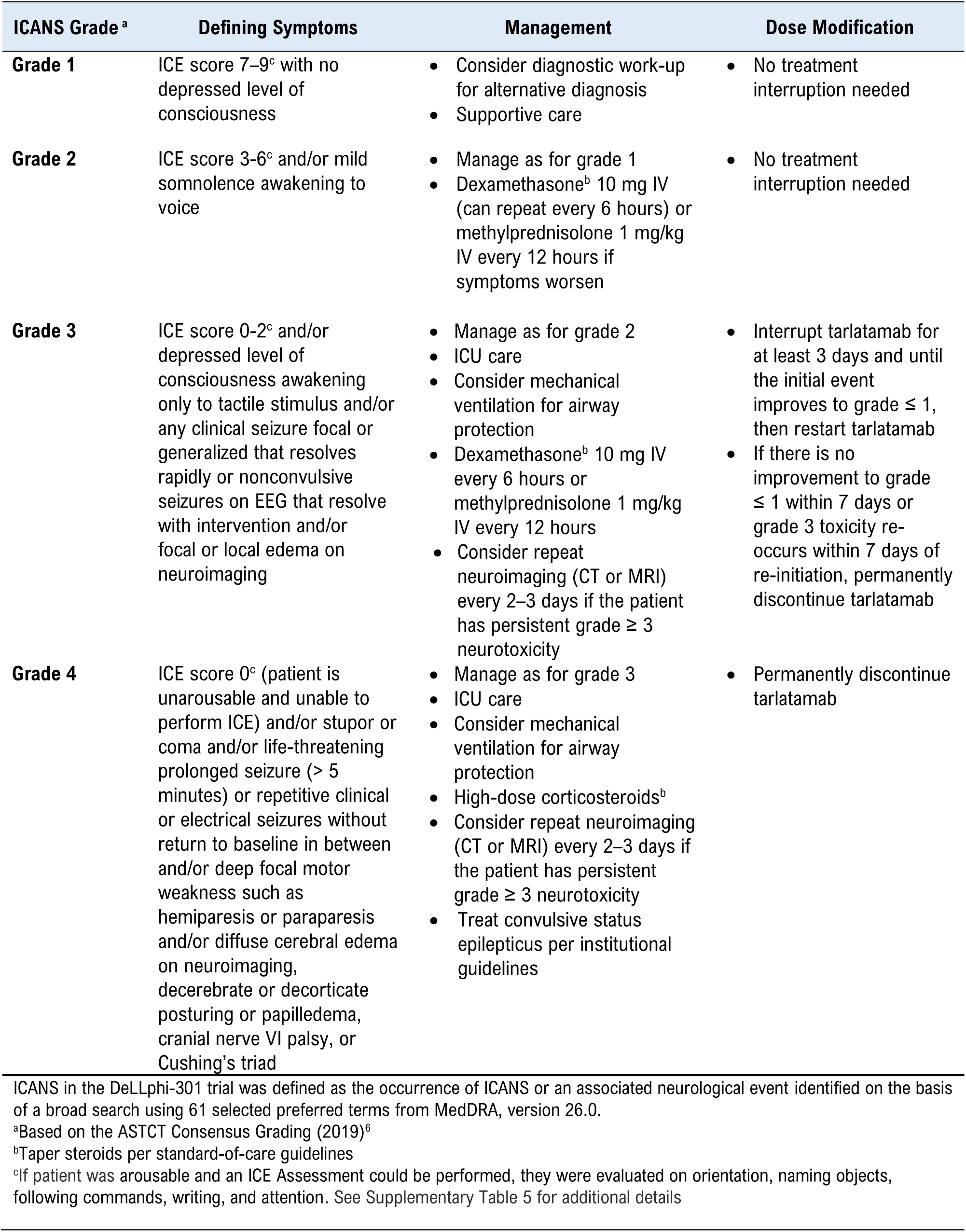
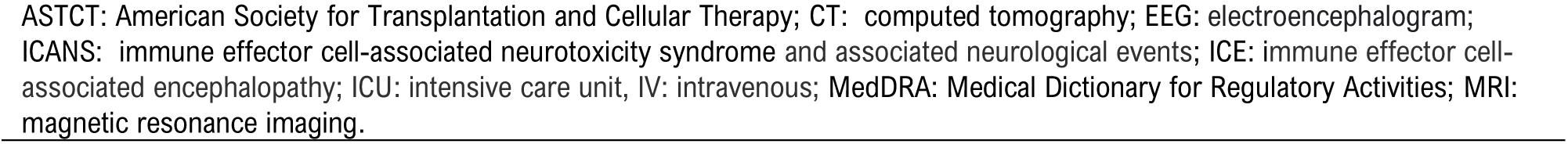
ICANS grade, defining symptoms, management, and dose modification as used in the DeLLphi-301 trial.

| ICANS Grade <sup>a</sup> | Defining Symptoms | Management | Dose Modification |
| --- | --- | --- | --- |
| **Grade 1** | ICE score 7–9<sup>c</sup> with no depressed level of consciousness | Consider diagnostic work-up for alternative diagnosis<br>Supportive care | No treatment interruption needed |
| **Grade 2** | ICE score 3–6<sup>c</sup> and/or mild somnolence awakening to voice | Manage as for grade 1<br>Dexamethasone<sup>b</sup> 10 mg IV (can repeat every 6 hours) or methylprednisolone 1 mg/kg IV every 12 hours if symptoms worsen | No treatment interruption needed |
| **Grade 3** | ICE score 0–2<sup>c</sup> and/or depressed level of consciousness awakening only to tactile stimulus and/or any clinical seizure focal or generalized that resolves rapidly or nonconvulsive seizures on EEG that resolve with intervention and/or focal or local edema on neuroimaging | Manage as for grade 2<br>ICU care<br>Consider mechanical ventilation for airway protection<br>Dexamethasone<sup>b</sup> 10 mg IV every 6 hours or methylprednisolone 1 mg/kg IV every 12 hours<br>Consider repeat neuroimaging (CT or MRI) every 2–3 days if the patient has persistent grade ≥ 3 neurotoxicity | Interrupt tarlatamab for at least 3 days and until the initial event improves to grade ≤ 1, then restart tarlatamab<br>If there is no improvement to grade ≤ 1 within 7 days or grade 3 toxicity re-occurs within 7 days of re-initiation, permanently discontinue tarlatamab |
| **Grade 4** | ICE score 0<sup>c</sup> (patient is unarousable and unable to perform ICE) and/or stupor or coma and/or life-threatening prolonged seizure (> 5 minutes) or repetitive clinical or electrical seizures without return to baseline in between and/or deep focal motor weakness such as hemiparesis or paraparesis and/or diffuse cerebral edema on neuroimaging, decerebrate or decorticate posturing or papilledema, cranial nerve VI palsy, or Cushing's triad | Manage as for grade 3<br>ICU care<br>Consider mechanical ventilation for airway protection<br>High-dose corticosteroids<sup>b</sup><br>Consider repeat neuroimaging (CT or MRI) every 2–3 days if the patient has persistent grade ≥ 3 neurotoxicity<br>Treat convulsive status epilepticus per institutional guidelines | Permanently discontinue tarlatamab |
| ASTCT: American Society for Transplantation and Cellular Therapy; CT: computed tomography; EEG: electroencephalogram;<br>ICANS: immune effector cell-associated neurotoxicity syndrome and associated neurological events; ICE: immune effector cell-associated encephalopathy; ICU: intensive care unit, IV: intravenous; MedDRA: Medical Dictionary for Regulatory Activities; MRI:<br>magnetic resonance imaging. |  |  |  |

### Strategies to mitigate the risk of developing CRS and ICANS used in the DeLLphi-301 trial

#### Pretreatment evaluation

Prior to treatment in the DeLLphi-301 trial, patients received physical and neurological examinations and laboratory testing that included a complete blood count, blood chemistry panel testing, and quantitation of C-reactive protein levels. Baseline hematological, renal, hepatic, pulmonary, and cardiac functions were also assessed prior to the first dose of tarlatamab.

#### Step-up dosing

Step-up dosing involves starting with a lower dose and escalating to the target dose in order to gradually prime the immune system and reduce the risk and/or severity of CRS.^19^ In the DeLLphi-301 trial, tarlatamab was administered as a step-up dose of 1 mg on C1D1, followed by 10 mg tarlatamab on C1D8 and C1D15. Thereafter, patients received 10 mg tarlatamab every 2 weeks until disease progression or unacceptable toxicity. If tarlatamab administration was delayed, the protocol defined restart guidelines for neurologic events, CRS, tumor lysis syndrome, non-febrile neutropenia, pituitary gland dysfunction, and hepatotoxicity. For all other treatment interruptions and delays, the protocol outlined restart guidelines as described in **Supplementary Table 8**.

#### Concomitant medications

Prophylactic administration of corticosteroids has been shown to lower the incidence and severity of CRS and ICANS.^12, 20, 21^ In addition, IV hydration may prevent an escalation in the severity of CRS by offsetting the effects of any potential hypotension.^22^ In the DeLLphi-301 trial, patients received dexamethasone 8 mg (or equivalent) IV within 1 hour prior to tarlatamab infusion on C1D1 and C1D8. Patients also received IV hydration (1 liter normal saline over 4–5 hours) immediately after all tarlatamab doses in cycle 1.

#### Post-infusion monitoring

For the dose-evaluation (part 1) and dose-expansion (part 2) parts of the DeLLphi 301 trial,^2^ all patients received their C1D1 and C1D8 doses in the hospital and remained hospitalized for 48 hours for observation and AE monitoring. Hospitalization was not required on C1D15 unless the patient had experienced any grade CRS or neurological events following the C1D1 or C1D8 doses; otherwise, the patient was observed for 8 hours following infusion **(Table 2)**. The frequency at which vital signs were assessed during hospitalization is shown in **Supplementary Table 9**. Hospitalization requirements from treatment cycle 2 onwards are detailed in **Table 2**.

In the part 3, reduced hospitalization sub-study, patients were monitored for 24 hours (either at an inpatient unit or outpatient infusion center) following the first two doses of tarlatamab (on C1D1 and C1D8). Vital signs were assessed as shown in **Supplementary Table 9**. Patients who experienced grade ≥ 2 CRS or grade ≥ 1 ICANS following C1D1 and C1D8 infusions had increased additional hospitalization requirements. Patients were observed for 6–8 hours if not hospitalized. CRS was graded and treated as described previously and as shown in **Table 4**. The safety profile in patients monitored for 24 hours post-infusion during cycle 1 was generally similar to that in patients monitored for 48 hours post-infusion. ^2^

### Post-hospitalization requirements for the reduced hospitalization substudy

Discharged patients were asked to stay within 1 hour of the infusion site for 48 hours and within 1 hour of a hospital for 72 hours after the C1D1 and C1D8 doses. A home companion was required for 72 hours following the C1D1 and C1D8 infusions. These considerations allowed for close monitoring, early detection of CRS, and prompt management if needed.

### Dysgeusia

The neuroendocrine transcription factor *ASCL1* drives the late-stage differentiation of certain types of taste bud cells.^23^ As *ASCL1* also regulates DLL3 expression,^24^ it can be speculated that these taste bud cells express DLL3. Tarlatamab may lead to T-cell-mediated destruction of these DLL3-expressing taste bud cells causing dysgeusia.

In the DeLLphi-301 trial, dysgeusia was reported in 32% of patients (42/133), with a median time to onset of 34 days (IQR, 21–51). (**Figure 1**).^23, 24^ Within the study protocol, no recommendations were provided on the management of dysgeusia. Some DeLLphi-301 investigators used previously published strategies for managing dysgeusia.^25^ These strategies include nutritional counseling from a registered dietitian and maintenance of good oral dental hygiene (thorough brushing of teeth at least twice a day). Other published strategies include maintenance of adequate hydration by drinking a minimum of 1.5–2 liters of energy-free beverages such as water or unsweetened tea per day, periodic chewing of cardamom, and inhaling the smell of cloves and lemon at least twice a day for about 15 seconds.^25^

### Neutropenia

Neutropenia is an identified risk for patients receiving tarlatamab.^26^ In the DeLLphi-301 trial, treatment-emergent neutropenia was reported in 16% of patients (21/133). Based on 21 patients with events, the median time to onset of neutropenia was 56 days (IQR, 30–86) (**Figure 1**). Febrile neutropenia was reported in less than 1% of patients (1/133). Neither neutropenia nor febrile neutropenia led to tarlatamab discontinuation. In the clinical trial setting, institution-specific protocols were suggested for the management of grade 1 or 2 neutropenia. For grade ≥ 3 neutropenia, treatment interruption was suggested until the severity decreased to grade 2 or lower. The use of granulocyte colony-stimulating factor (G-CSF) was permitted for the management of grade 3 neutropenia. Tarlatamab discontinuation was advised if an initial grade 3 non-febrile neutropenia did not improve to grade 1 or lower within 3 weeks or if an initial grade 4 non-febrile neutropenia lasted for > 7 days or re-occurred at a lower dose level.

### Liver enzyme elevations

Alanine aminotransferase (ALT) increase was observed in 15 patients (11.3%) with two patients (1.5%) experiencing a grade 3 event. The median time to onset of ALT increase from the first tarlatamab dose was 29 days (IQR, 7–113). For the 14 patients (10.5%) with increased aspartate aminotransferase (AST) levels, the median time to onset was 51 days (IQR, 3–196). One patient had elevated aminotransferase and total bilirubin > 2 x upper limit of normal (ULN), and alkaline phosphatase < 2 x ULN. Elevated blood bilirubin was observed in four patients (3%). No patient reported tarlatamab discontinuation due to increased ALT or AST. The changes in liver enzyme and bilirubin levels were generally transient, with no evidence suggesting extensive hepatocellular damage. Documenting baseline levels of liver enzymes and bilirubin before the start of tarlatamab and monitoring before each dose of tarlatamab and as clinically indicated was performed in the DeLLphi-301 trial.

### Other TEAEs

Other AEs reported in DeLLphi-301 included decreased appetite, fatigue, anemia and infections. Decreased appetite and fatigue are AEs that, if not appropriately monitored and managed, can negatively impact long-term patient experience. In DeLLphi-301, decreased appetite was reported in 36% (48/133) of patients and fatigue in 24% (32/133) of patients. Standard nonpharmacologic approaches such as encouraging patients to undertake an optimal level of activity under the guidance of a health care provider, massage therapy, acupuncture, psychosocial interventions, nutrition consultations, and pharmacologic interventions have been suggested for the management of fatigue in patients undergoing cancer treatment.^27^ Anemia was observed in 40 patients (30%) with a median time to onset of 13.0 days (range, 1–275) from the first tarlatamab dose. (**Figure 1**).

Among 57 patients with infections, the median time to onset of 37.0 days (range, 2–436). Grade ≥ 3 treatment-emergent infections, including opportunistic infections, occurred in 13% of patients (17/133). The most frequent grade ≥ 3 treatment-emergent infections were pneumonia (n = 6), respiratory tract infection (n = 3), device-related infection, and pneumonia aspiration (n = 2 each). Infections were managed using relevant institution-specific protocols. For all infections, the prescribing information recommends withholding tarlatamab until resolution, or in the case of a grade 3 infection (graded per Common Terminology Criteria for Adverse Events [CTCAE] v5.0 guidelines^17^), withholding tarlatamab until infection improved to grade 1 or lower.^28^ Permanent discontinuation of tarlatamab was recommended in the case of a grade 4 infection.^28^

### Considerations for Patient Management

Early identification of the signs and symptoms of AEs and an awareness of management practices may help health care providers optimize care for patients receiving tarlatamab. As such, patients in the DeLLphi-301 trial were monitored in a healthcare facility for 24–48 hours following the first two doses of tarlatamab. Investigators also took into consideration the ability to rapidly transfer patients to an intensive care unit (ICU) to manage higher grade CRS or ICANS events. Following hospital discharge, patients were to be accompanied by a caregiver and remain within 1 hour of an appropriate healthcare setting for a total of 72 hours after completion of the infusion following the first two tarlatamab doses. Other considerations identified by investigators that can aid in optimizing the management of CRS and ICANS are listed in **Table 7**. Healthcare providers educated patients and caregivers on recognizing the signs and symptoms of CRS and ICANS. They were taught to recognize CRS symptoms such as fever, difficulty breathing or unusual shortness of breath, headache, chest pain, palpitations, persistent nausea or vomiting, chills, and/or myalgia and advised on the appropriate administration of acetaminophen. Patients and caregivers were also instructed on monitoring changes in vital signs (body temperature [≥ 38°C], blood pressure, oxygen saturation, and heart rate) for early identification of AEs. When a patient living in a rural setting developed symptoms suggestive of CRS, the patient and caregiver were instructed to treat with dexamethasone and visit the emergency room (ER) for further help.

**Table 7:**
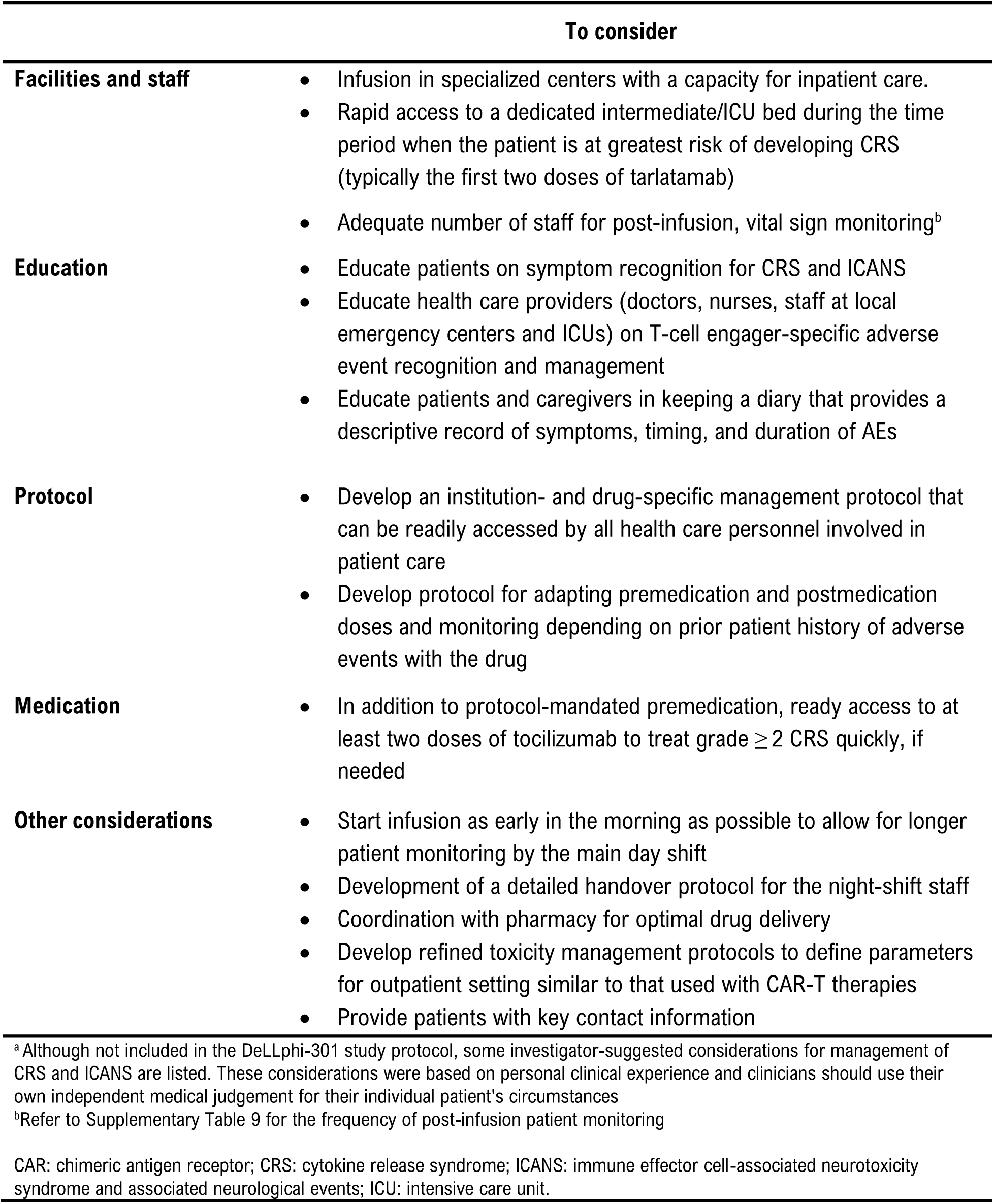
Investigator-suggested considerations for optimal management of CRS and ICANS^a^.

Patients and caregivers were also taught to recognize neurological symptoms that could be manifestations of ICANS, such as headache, dysgeusia, dizziness, insomnia, muscular weakness, ataxia, amnesia, confusion, go to the ER if they experienced any neurologic symptoms at any time during their treatment with tarlatamab. Patients were also advised to refrain from driving, operating heavy or potentially dangerous machinery, and engaging in hazardous occupations or activities until their neurologic symptoms resolved.

Patient or caregiver diaries with a descriptive record of symptoms, timing, and duration of AEs can assist healthcare providers with the management of AEs (**Table 7**). Additionally, at one site, patients and caregivers were provided wallet cards with key contact information and asked to show the wallet card to healthcare providers.

## Discussion

To date, the experience with CAR-T-cell therapies in hematological malignancies has driven the management of CRS and ICANS. However, it is important to distinguish the safety profile of tarlatamab. In the DeLLphi-301 trial, CRS and ICANS was generally low-grade and manageable with standard supportive care that included acetaminophen, IV hydration, and/or glucocorticoids. CRS-associated fever usually occurred within hours of tarlatamab administration and was sometimes the only symptom before it resolved. Progression to higher-grade CRS generally took hours and was, in most instances, managed conservatively. ICANS occurred with a relatively low incidence, was low-grade, and was typically managed with corticosteroids. While ICANS occurred more frequently in patients with treated, stable brain metastases in the DeLLphi-301 phase 2 trial, this correlation was not observed in the DeLLphi-300 phase 1 trial.^29, 30^

Additional AEs experienced by patients treated with tarlatamab include cytopenias, liver enzyme elevations and dysgeusia, which typically can be managed with standard of care **(Figure 3).**

**Figure 3:**
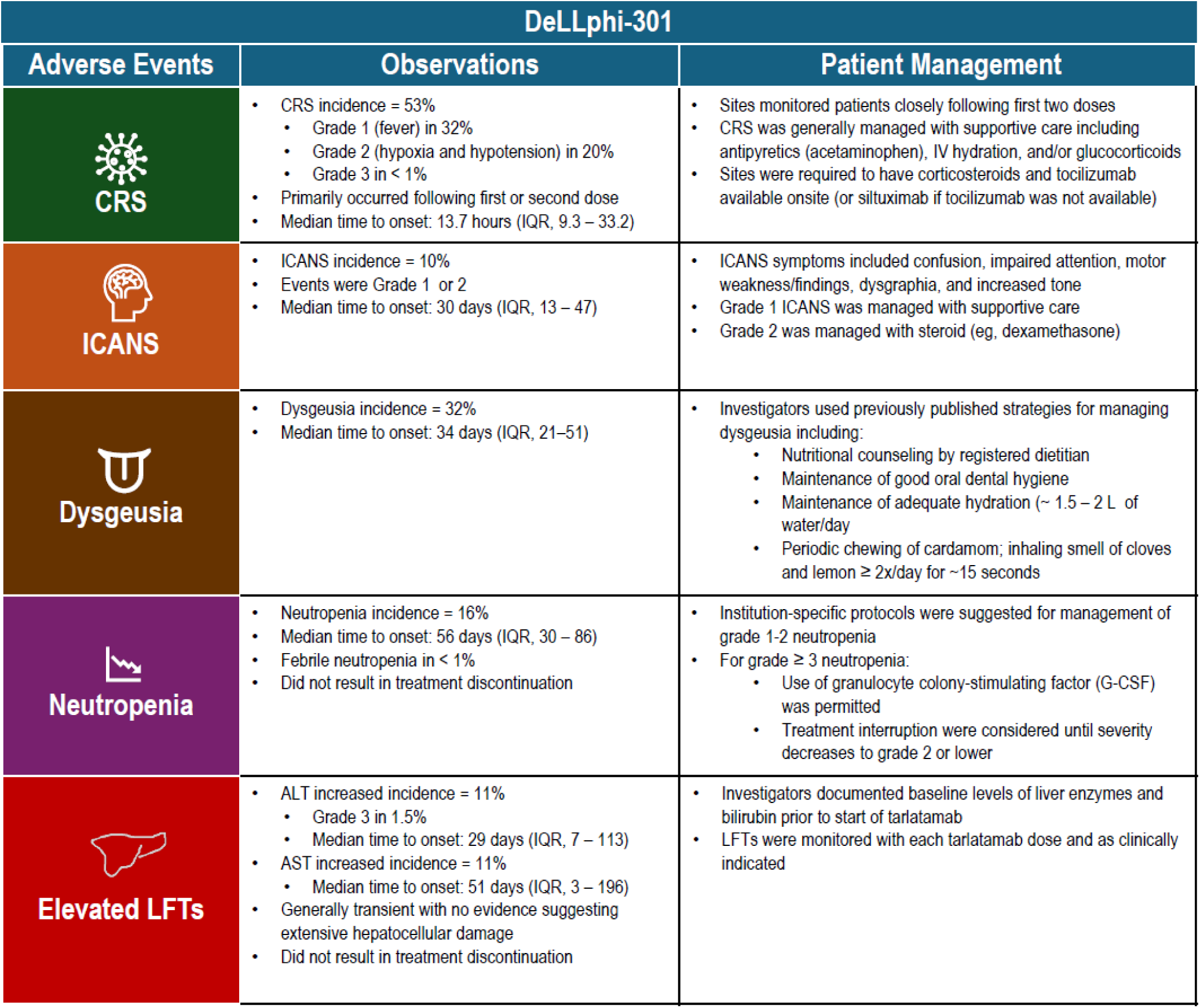
Common and Adverse Events of Interest in the DeLLphi-301 Trial Study.

## Conclusions

Clinical trial experience indicates that tarlatamab has a manageable safety profile with a low discontinuation rate (3%) due to treatment-related AEs. Tarlatamab-associated AEs, including immune-related AEs such as CRS and ICANS were primarily grade 1 or 2 in severity and mostly occurred early during treatment (cycle 1) and could generally be managed with standard supportive care. Monitoring, detection, and appropriate management of AEs, such as CRS and ICANS will remain important for maintaining the continuity of care. Knowledge of other AEs such as neutropenia, transient liver enzyme elevations, and dysgeusia is also important to consider, especially during the long-term treatment of patients on tarlatamab. Continuous patient education by nurses and pharmacists may help with symptom monitoring and management and optimize long-term patient treatment experience.

## Acknowledgments

Medical writing support, including development of a draft outline and subsequent drafts in consultation with the authors, assembling tables and figures, collating author comments, copyediting, fact checking, and referencing, was provided by Sukanya Raghuraman, PhD of Cactus Life Sciences (part of Cactus Communications), Martha Mutomba, PhD (on behalf of Amgen), and Dawn Nicewarner, PhD, CMPP (Amgen).

## Role of the funding source

Amgen funded medical writing support, paid publication costs, and paid the open access charge for this article. With the exception of Shuang Huang, Ali Hamidi, and Sujoy Mukherjee (Amgen employees), Amgen had no involvement in the decision to submit the article for publication; this decision was made solely by the author group.

## Author contributions

Conceptualization: all authors. Writing -Review & Editing: all authors. All authors meet the International Committee of Medical Journal Editors criteria for authorship of this article, take responsibility for the integrity of the work as a whole, and have given their approval for publication of the final version.

## Ethics statement

The DeLLphi-301 trial (NCT05060016) was conducted in accordance with the International Council for Harmonisation Good Clinical Practice guidelines and the principles of the Declaration of Helsinki. The protocol and amendments were approved by the institutional review board at each participating site and by regulatory authorities in the participating countries as listed: **Austria:** Ethikkommission fuer das Bundesland Salzburg, Ethikkommission des Landes Niederoesterreich; **Belgium:** Ethische Commissie Onderzoek UZ/KU Leuven, Universitair Ziekenhuis Gent -Ethisch Comite; **Denmark:** Region Hovedstaden; **France:** Comite de Protection des Personnes Ile de France VI -Hopital Pitie-Salpetriere; **Germany:** Ethikkommission bei der Aerztekammer Schleswig-Holstein, Ethik-Kommission der Medizinischen Fakultaet der Universitaet Wuerzburg, Ethikkommission der Universitaet Koeln; **Greece:** National Ethics Committee; **Italy:** Comitato Etico Territoriale Lazio Area 5, Comitato Etico Sezione Istituti Fisioterapici Ospitalieri, Comitato Etico Unico per la Provincia di Parma; **Japan:** National Cancer Center Institutional Review Board 2-J, Wakayama Medical University Institutional Review Board, Shizuoka Cancer Center Institutional Review Board, Kindai University Hospital Institutional Review Board, IRB of Okayama University Hospital, Aichi Cancer Center Institutional Review Board, The Cancer Institute Hospital of Japanese Foundation for Cancer Research Institutional Review Board; **Netherlands:** Stichting Beoordeling Ethiek Biomedisch Onderzoek, Centrale Commissie Mensgebonden Onderzoek; **Poland:** Niezalezna Komisja Bioetyczna do spraw Badan Naukowych przy Gdanskim Uniwersytecie Medycznym, Naczelna Komisja Bioetyczna; **Portugal:** Comissao de Etica para a Investigacao Clinica; **Republic of Korea:** Asan Medical Center Institutional Review Board, Samsung Medical Center Institutional Review Board, Yonsei University Health System Severance Hospital Institutional Review Board, National Cancer Center IRB, Helsingin ja Uudenmaan Sairaanhoitopiiri tutkimuseettiset toimikunnat, The Catholic University of Korea Seoul St Marys Hospital Institutional Review Board; **Spain:** Instituto de Investigacion Hospital 12 de Octubre i12; **Switzerland:** Commission Cantonale d’etique de la Recherche, Kantonale Ethikkommission Zuerich; **Taiwan:** Taipei Veterans General Hospital Institutional Review Board; **United Kingdom:** North East -Tyne and Wear South Research Ethics Committee; **United States:** Western Institutional Review Board Copernicus Group, University Hospitals Cleveland Medical Center Institutional Review Board, Dana-Farber Cancer Institute Institutional Review Board, Advarra IRB, Advocate Health -Wake Forest University Health Sciences Institutional Review Board, University of Arkansas for Medical Sciences IRB. All patients provided written informed consent.

## Conflicts of interest

**Jacob Sands** reports honoraria from Pfizer; consulting or advisory fees from AstraZeneca, Medtronic, Daiichi Sankyo/UCB Japan, Sanofi, Boehringer Ingelheim, PharmaMar, Guardant Health, AbbVie, Gilead Sciences, Lilly, G1 Therapeutics; research funding from Amgen and Harpoon, and travel/ accommodation/ and expenses support from AstraZeneca.

**Stéphane Champiat** reports honoraria from Amgen, AstraZeneca, Bristol Myers Squibb, MSD, Novartis, Roche, Fresenius Kabi, Eisai Europe, Genmab, Janssen, Merck KGaA, Merck Serono, Astellas Pharma, SERVIER and Takeda; consulting or advisory fees from Alderaan Biotechnology, Amgen, AstraZeneca, Avacta Life Sciences, Celanese, Domain Therapeutics, Ellipses Pharma, Genmab, Immunicom, Nanobiotix, Oncovita, Pierre Fabre, Seagen, Tatum Bioscience, Tollys, UltraHuman8, NextCure, BioNTech SE, Mariana Oncology, Takeda, BeiGene; research funding (to institution) from AstraZeneca, Bristol Myers Squibb, Boehringer Ingelheim, Janssen-Cilag, Merck, Novartis, Pfizer, Roche (Inst, Sanofi, Abbvie, Adaptimmune, Aduro Biotech, Agios, Amgen, arGEN-X BVBA, Arno Therapeutics, Astex Pharmaceuticals, AstraZeneca, Bayer, BB Biotech Ventures, BeiGene, BioAlliance Pharma, BioNTech, Blueprint Medicines, Boehringer Ingelheim, Boston Pharmaceuticals, Bristol Myers Squibb, Celgene, Cephalon, Chugai Pharma, Clovis Oncology, Cullinan Oncology, Daiichi Sankyo, Debiopharm Group, Eisai, Lilly, Exelixis, FORMA Therapeutics, GamaMabs Pharma, Genentech, Gilead Sciences, GlaxoSmithKline, Glenmark, H3 Biomedicine, Roche, Incyte, Innate Pharma, Pierre Fabre, SERVIER, Janssen-Cilag, Kura Oncology, Kyowa Hakko Kirin, Loxo, Lytix Biopharma, MedImmune, Menarini, Merck KGaA, Merck Sharp & Dohme, Merrimack, Merus, Millennium, Molecular Partners, Nanobiotix, Nektar, Nerviano Medical Sciences, Novartis, Octimet, Oncoethix, OncoMed, Oncopeptides, Onyx, Orion, Oryzon Genomics, Ose Pharma, Pfizer, PharmaMar, Philogen, Pierre Fabre, Plexxikon, RigonTEC, Sanofi/Aventis, Sierra Oncology, Sotio, Syros Pharmaceuticals, Taiho Pharmaceutical, Tesaro, Tioma Therapeutics, Wyeth, Xencor, Y’s Therapeutics, Cytovation, Eisai/H3 Biomedicine, ImCheck therapeutics, Molecular Partners, MSD, OSE Immunotherapeutics, Pierre Fabre, Sanofi, Sotio, Transgene, Boehringer Ingelheim, Abbvie, Amgen, Adlai Nortye, AVEO, Basilea Pharmaceutical, BBB Technologies, Bicycle Therapeutics, CASI Pharmaceuticals, CellCentric, CureVac, Faron Pharmaceuticals, ITeos Therapeutics, Relay Therapeutics, Seagen, Transgene, Turning Point Therapeutics, GlaxoSmithKline, GlaxoSmithKline, Immunocore, Replimune, Roche/Genentech, Seagen, Bolt Biotherapeutics, Centessa Pharmaceuticals, Veracyte; patents, royalties, other intellectual property for patent number WO2010039223A2 (T-cell immunogens derived from anti-viral proteins and methods of using same); travel, accommodations, and expenses support from MSD, AstraZeneca, Amgen, Bristol Myers Squibb, Merck, OSE Immunotherapeutics, Roche, Sotio, Boehringer Ingelheim; other relationships with AstraZeneca (Institutional), Bayer (Institutional), Bristol Myers Squibb (Institutional), Boehringer Ingelheim (Institutional), Boehringer Ingelheim (Institutional), Johnson & Johnson (Institutional), Lilly (Institutional), MedImmune (Institutional), Merck (Institutional), Pfizer (Institutional), Roche (Institutional), Roche (Institutional), GlaxoSmithKline (Institutional), NH TherAguix (Institutional).

**Horst-Dieter Hummel** reports honoraria from Amgen, Boehringer Ingelheim,Bristol Myers Squibb/Pfizer, Amgen (Institutional), Revolution Medicines (Institutional), Merck (Institutional), Bristol Myers Squibb/Pfizer (Institutional), AstraZeneca (Institutional), and Johnson & Johnson/Janssen; consulting or advisory role fees from Amgen, Boehringer Ingelheim; travel/accommodations/expenses support from Amgen, Boehringer Ingelheim

**Hossein Borghaei** reports stock and other ownership interests in Sonnet, Rgenix, Nucleai; honoraria from Bristol Myers Squibb, Celgene, Axiom Biotechnologies, Pfizer, Amgen, Regeneron, Daiichi Sankyo/UCB Japan; consulting or advisory role fees from Bristol Myers Squibb, Lilly, Genentech, Pfizer, Boehringer Ingelheim, EMD Serono, Novartis, Merck, AstraZeneca, Genmab, Regeneron, BioNTech, Abbvie, PharmaMar, Takeda, Amgen, Sonnet, Rgenix, Beigene, Jazz Pharmaceuticals, Mirati Therapeutics, Guardant Health, Janssen Oncology, ITeos Therapeutics, Natera, oncocyte, Puma Biotechnology, BerGenBio, Bayer, Bayer, IO Biotech, RAPT Therapeutics, Grid Therapeutics; research funding to the institution from Bristol Myers Squibb, Lilly, Amgen; travel/accommodations/expenses support from Bristol Myers Squibb, Lilly, Clovis Oncology, Celgene, Genentech, Novartis, Merck, Amgen, EMD Serono, Regeneron, Mirati Therapeutics; other relationships with University of Pennsylvania, Takeda, Incyte, Novartis, and SpringWorks Therapeutics.

**David Paul Carbone** is employed by James Cancer Center; reports honoraria from AstraZeneca and Bristol-Myers Squibb-Ono Pharmaceutical; consulting or advisory fees from Abbvie, Arcus Biosciences, AstraZeneca, BMS Israel, Bristol-Myers Squibb, Curio Science, EMD Serono, Genentech, Genentech/Roche, GlaxoSmithKline, Intellisphere, InThought, Iovance Biotherapeutics, Janssen, Jazz Pharmaceuticals, JNJ, Johnson & Johnson/Janssen, Lilly, Merck, Merck KGaA, Mirati Therapeutics, MSD Oncology, NCCN/AstraZeneca, Novartis, Novocure, OncLive/MJH Life Sciences, Oncohost, Pfizer, PPD, Regeneron, Roche, Roche/Genentech, and Sanofi.

**Jennifer Carlisle** reports personal fees for participation in advisory boards from Sanofi, Amgen, and Novocure; research funding to institution from AstraZeneca, Amgen, Hutchmed and Parexel.

**Noura J. Choudhury** reports consulting or advisory fees from Abbvie, SG1 Therapeutics, Harpoon therapeutics, and Sanofi; research funding from Abbvie (Institutional), Amgen (Institutional), Harpoon therapeutics (Institutional), Merck (Institutional), Monte Rosa Therapeutics (Institutional), and Roche/Genentech (Institutional); and royalties from Wolters Kluwer (Pocket Oncology).

**Jeffrey M. Clarke** reports grants and personal fees from Bristol Myers Squibb, personal fees from CDR Life, Coherus, and Black Diamond; grants and personal fees from AstraZeneca, Moderna, Merck, and Amgen; personal fees from G1, Sanofi, and Corbus; grants from Grid, Abel Zeta, and Adaptimmune; grants and personal fees from Spectrum; grants from Genentech; and personal fees from DSI and BioThera outside the submitted work.

**Shirish M. Gadgeel** reports honoraria from Merck; consulting or advisory fees from Genentech/Roche, AstraZeneca, Bristol Myers Squibb, Takeda, Daichii-Sanyko, Novartis, Blueprint Medicines, Lilly, Pfizer, Janssen Oncology, Mirati Therapeutics, Merck, Eisai, Gilead Sciences, GlaxoSmithKline, Arcus Biosciences, Bayer, and Regeneron; research funding from Merck, Genentech/Roche (Institutional), Merck (Institutional), Blueprint Medicines (Institutional), Astellas Pharma (Institutional), Daiichi Sankyo (Institutional), I-Mab (Institutional), Nektar (Institutional), AstraZeneca, AstraZeneca (Institutional), Pfizer (Institutional), Amgen (Institutional), Turning Point Therapeutics (Institutional), Regeneron (Institutional), Mirati Therapeutics (Institutional), Nektar (Institutional), Janssen Oncology (Institutional), BioMed Valley Discoveries (Institutional), Ymabs Therapeutics Inc (Institutional), Calithera Biosciences (Institutional), InventisBio (Institutional), Daichii Sanyko (Institutional), Dragonfly Therapeutics (Institutional), eFFECTOR Therapeutics (Institutional), Elevation Oncology (Institutional), Erasca, Inc (Institutional), Helsinn Therapeutics (Institutional), Incyte (Institutional), Numab (Institutional), Verastem (Institutional), Regeneron (Institutional), and Debiopharm Group (Institutional); travel/accommodations/expenses support from Mirati Therapeutics; and other Relationships with AstraZeneca.

**Hiroki Izumi** reports honoraria from Bristol-Myers Squibb Japan, Chugai/Roche, Lilly, Merck, MSD, Ono Pharmaceutical, and Takeda; consulting or advisory fees from Amgen; research funding from Abbvie (Institutional), Amgen (Institutional), ArriVent Biopharma (Institutional), AstraZeneca (Institutional), Eisai/MSD (Institutional), Ono Pharmaceutical (Institutional), and Takeda (Institutional).

**Alejandro Navarro** reports consulting or advisory fees from Adium Pharma, Amgen, Boehringer Ingelheim, Bristol Myers Squibb Foundation, Eczacibasi, Pfizer, and Takeda; speakers’ bureau fees from AstraZeneca Spain, and Roche; expert testimony fees from Hengenix, Medsir, and Oryzon Genomics; travel/accommodations/expenses support from Boehringer Ingelheim, Pfizer, and Roche.

**Philip E. Lammers** reports consulting or advisory fees from AstraZeneca, Daiichi Sankyo, Merck, Pfizer, Roche/Genentech, Sanofi, and Teva.

**Shuang Huang** is employed by Amgen and owns stock and/or stock options in Amgen.

**Ali Hamidi** is employed by Amgen and owns stock and/or stock options in Amgen.

**Sujoy Mukherjee** is employed by Amgen and owns stock and/or stock options in Amgen.

**Taofeek K. Owonikoko** owns stock and has other ownership interests in Cambium Medical Technologies, Coherus Biosciences, GenCart, and Taobob LLC; reports consulting or advisory fees from Abbvie, Amgen, AstraZeneca, Bayer, BeiGene, BerGenBio, Boehringer Ingelheim, Bristol-Myers Squibb, Celgene, Coherus Biosciences, Daichi, Eisai, Eisai, EMD Serono, Exelixis, G1 Therapeutics, Heat Biologics, Ipsen, Janssen, Jazz Pharmaceuticals, Lilly, MedImmune, Merck, Meryx Pharmaceuticals, Novartis, PharmaMar, Puma Biotechnology, Roche/Genentech, Takeda, Triptych Health Partners, and Xcovery; reports research funding from Abbvie (Institutional), Amgen (Institutional), AstraZeneca/MedImmune (Institutional), Bayer (Institutional), Boehringer Ingelheim (Institutional), Bristol-Myers Squibb (Institutional), Calithera Biosciences (Institutional), Corvus Pharmaceuticals, G1 Therapeutics (Institutional), GlaxoSmithKline (Institutional), Incyte (Institutional), Loxo/Lilly (Institutional), Merck (Institutional), Meryx Pharmaceuticals (Institutional), Novartis (Institutional), Oncorus (Institutional), Pfizer (Institutional), Regeneron (Institutional), and Roche/Genentech (Institutional); has patents, royalties, and other intellectual property related to: DR4 Modulation and its Implications in EGFR-Target Cancer Therapy Ref:18089 PROV (CSP) United States Patent Application No. 62/670,210 June 26, 2018 (Co-Inventor) (Institutional); Overcoming Acquired Resistance to Chemotherapy Treatments Through Suppression of STAT3 (Institutional); Selective Chemotherapy Treatments and Diagnostic Methods Related Thereto (Institutional); and Soluble FAS Ligand as a Biomarker of Recurrence in Thyroid Cancer; provisional patent 61/727,519 (Inventor) (Institutional); reports travel/accommodations/expenses support from AstraZeneca and Janssen; other relationships with EMD Serono, Novartis, and Roche/Genentech; and an uncompensated relationship with Reflexion Medical.

**The remaining authors** have no disclosures.

## Data availability statement

Qualified researchers may request data from Amgen clinical studies. Details are available at https://www.amgen.com/datasharing

## Funding statement

This study was funded by Amgen Inc.

## Notes

### Clinical Trial

NCT05060016

### Clinical Protocols

https://www.nejm.org/doi/full/10.1056/NEJMoa2307980#supplementary-materials

### Author Declarations

The DeLLphi-301 trial (NCT05060016) was conducted in accordance with the International Council for Harmonisation Good Clinical Practice guidelines and the principles of the Declaration of Helsinki. The protocol and amendments were approved by the institutional review board at each participating site and by regulatory authorities in the participating countries as listed: Austria: Ethikkommission fuer das Bundesland Salzburg, Ethikkommission des Landes Niederoesterreich; Belgium: Ethische Commissie Onderzoek UZ/KU Leuven, Universitair Ziekenhuis Gent - Ethisch Comite; Denmark: Region Hovedstaden; France: Comite de Protection des Personnes Ile de France VI - Hopital Pitie-Salpetriere; Germany: Ethikkommission bei der Aerztekammer Schleswig-Holstein, Ethik-Kommission der Medizinischen Fakultaet der Universitaet Wuerzburg, Ethikkommission der Universitaet Koeln; Greece: National Ethics Committee; Italy: Comitato Etico Territoriale Lazio Area 5, Comitato Etico Sezione Istituti Fisioterapici Ospitalieri, Comitato Etico Unico per la Provincia di Parma; Japan: National Cancer Center Institutional Review Board 2-J, Wakayama Medical University Institutional Review Board, Shizuoka Cancer Center Institutional Review Board, Kindai University Hospital Institutional Review Board, IRB of Okayama University Hospital, Aichi Cancer Center Institutional Review Board, The Cancer Institute Hospital of Japanese Foundation for Cancer Research Institutional Review Board; Netherlands: Stichting Beoordeling Ethiek Biomedisch Onderzoek, Centrale Commissie Mensgebonden Onderzoek; Poland: Niezalezna Komisja Bioetyczna do spraw Badan Naukowych przy Gdanskim Uniwersytecie Medycznym, Naczelna Komisja Bioetyczna; Portugal: Comissao de Etica para a Investigacao Clinica; Republic of Korea: Asan Medical Center Institutional Review Board, Samsung Medical Center Institutional Review Board, Yonsei University Health System Severance Hospital Institutional Review Board, National Cancer Center IRB, Helsingin ja Uudenmaan Sairaanhoitopiiri tutkimuseettiset toimikunnat, The Catholic University of Korea Seoul St Marys Hospital Institutional Review Board; Spain: Instituto de Investigacion Hospital 12 de Octubre i12; Switzerland: Commission Cantonale d'etique de la Recherche, Kantonale Ethikkommission Zuerich; Taiwan: Taipei Veterans General Hospital Institutional Review Board; United Kingdom: North East - Tyne and Wear South Research Ethics Committee; United States: Western Institutional Review Board Copernicus Group, University Hospitals Cleveland Medical Center Institutional Review Board, Dana-Farber Cancer Institute Institutional Review Board, Advarra IRB, Advocate Health - Wake Forest University Health Sciences Institutional Review Board, University of Arkansas for Medical Sciences IRB. All patients provided written informed consent.

### Summary of Updates

In 'CRS management in the DeLLphi-301 trial' section, noted that siltuximab was not used. Added paraneoplastic syndromes to DDx for ICANS. Added Supplementary Table 8 outlining specific restart guidelines. Clarified that protocol did not provide recommendation on management of dysgeusia.

