## Supplementary Material for "Practical Management of Adverse Events in Patients Receiving Tarlatamab, a DLL3-targeted Bispecific T-cell Engager Immunotherapy, for Previously Treated Small Cell Lung Cancer"

**Supplementary Table 1. Incidence of most frequent (≥ 20%) any-grade treatment-emergent adverse events with tarlatamab 10 mg in the DeLLphi-301 trial**

| **Preferred term for adverse event** | **Tarlatamab 10 mg**  **(N = 133)**  **n (%)** |
| --- | --- |
| ***Number of patients reporting treatment-emergent adverse events*** | ***133 (100)*** |
| Cytokine release syndrome | 70 (53) |
| Pyrexia | 50 (38) |
| Decreased appetite | 48 (36) |
| Dysgeusia | 42 (32) |
| Anemia | 40 (30) |
| Constipation | 39 (29) |
| Asthenia | 33 (25) |
| Fatigue | 32 (24) |

The safety analysis set included all patients who had received at least one dose of tarlatamab.

Adverse events were coded using MedDRA v26.1.

Events of small cell lung cancer/disease progression were excluded.

MedDRA: Medical Dictionary for Regulatory Activities.

**Supplementary Table 2.** **Incidence of common (> 2%) grade 3 or higher TEAEs in the tarlatamab 10 mg dose group in the DeLLphi-301 trial**

| **Preferred Term for Adverse Event** | **Tarlatamab 10 mg (N = 133) n (%)** |
| --- | --- |
| ***Number of patients reporting grade 3 or higher TEAEs*** | ***84 (63.2)*** |
| Lymphopenia^a^ | 18 (13.5) |
| Anemia | 10 (7.5) |
| Hyponatremia | 8 (6.0) |
| Neutropenia^b^ | 8 (6.0) |
| Asthenia | 7 (5.3) |
| Fatigue | 7 (5.3) |
| Pneumonia | 6 (4.5) |
| Decreased appetite | 4 (3.0) |
| Hypertension | 4 (3.0) |
| General physical health deterioration | 3 (2.3) |
| Hypokalemia | 3 (2.3) |
| Nausea | 3 (2.3) |
| Respiratory tract infection | 3 (2.3) |
| The safety analysis set includes all patients who received at least 1 dose of tarlatamab.  Adverse events were coded using MedDRA v26.1 and graded using CTCAE v5.0.  Events of small cell lung cancer/disease progression were excluded.  ^a^Adverse events for MedDRA-preferred terms “lymphopenia” and “lymphocyte count decreased.”  ^b^Adverse events for MedDRA-preferred terms “neutropenia” and “neutrophil count decreased.”  CTCAE: Common Terminology Criteria for Adverse Events; MedDRA: Medical Dictionary for Regulatory Activities; TEAE: treatment-emergent adverse event | |

**Supplementary Table 3. Other organ dysfunctions associated with CRS**

| **Organ System** | **Combination of CRS Signs and Symptoms** |
| --- | --- |
| Constitutional | Fever and/or rigors, malaise, fatigue, anorexia, myalgias, arthralgias,  nausea, vomiting, headache |
| Skin | Rash |
| Gastrointestinal | Nausea, vomiting, diarrhea |
| Respiratory | Tachypnea, hypoxemia |
| Cardiovascular | Tachycardia, widened pulse pressure, hypotension, increased cardiac output (early), potentially diminished cardiac output (late) |
| Coagulation | Elevated D-dimer, hypofibrinogenemia and/or bleeding |
| Renal | Azotemia |
| Hepatic | Transaminitis, hyperbilirubinemia |
| CRS, cytokine release syndrome. | |

**Supplementary Table 4. Interventions used for the management of CRS in DeLLphi-301**

|  | **Tarlatamab**  **10 mg**  **(N = 133) n (%)** |
| --- | --- |
| **Number of patients with at least one CRS event** | **70 (52.6)** |
| **Utilization of any CRS intervention listed below** | 22 (31.4) |
| Tocilizumab use | 8 (11.4) |
| Vasopressor use^a^  (excluding vasopressin) | 1 (1.4) |
| IV fluid use | 9 (12.9) |
| Low-flow (≤ 6 L/Min) supplemental oxygen use | 10 (14.3) |
| High-flow (> 6 L/min) supplemental oxygen use | 1 (1.4) |
| ^a^Patient required the use of a single vasopressor only.  CRS: cytokine release syndrome; IV: intravenous; N: Number of patients in the analysis set; n: number of patients with observed data. | |

**Supplementary Table 5. ASTCT ICANS consensus grading system for adults^6^**

| **Neurotoxicity Domain^a^** | **Grade 1** | **Grade 2** | **Grade 3** | **Grade 4** |
| --- | --- | --- | --- | --- |
| **ICE score^b^** | 7–9 | 3–6 | 0–2 | 0 (patient is unarousable and unable to perform ICE) |
| **Depression level of consciousness^c^** | Awakens spontaneously | Awakens to voice | Awakens only to tactile stimulus | Patient is unarousable or requires vigorous or repetitive tactile stimuli to arouse;  stupor or coma |
| **Seizure** | N/A | N/A | Any clinical seizure focal or generalized that resolves rapidly or nonconvulsive seizures on EEG that resolve with intervention | Life-threatening prolonged seizure (> 5 min); or repetitive clinical or electrical seizures without return to baseline in between |
| **Motor findings** | N/A | N/A | N/A | Deep focal motor weakness such as hemiparesis or paraparesis |
| **Elevated intracranial pressure /cerebral edema** | N/A | N/A | Focal/local edema on neuroimaging^d^ | Diffuse cerebral edema on neuroimaging; decerebrate or decorticate posturing; or cranial nerve VI palsy; or papilledema; or Cushing’s triad |
| ^a^Other signs and symptoms such as headache, tremor, myoclonus, asterixis, and hallucinations may occur and could be attributable to immune effector cell-engaging therapies. Although they are not included in this grading scale, careful attention and directed therapy may be warranted.  ^b^A patient with an ICE score of 0 may be classified as grade 3 ICANS if awake with global aphasia, but a  patient with an ICE score of 0 may be classified as grade 4 ICANS if unarousable.  ^c^Depressed level of consciousness should be attributable to no other cause (eg, no sedating medication).  ^d^Intracranial hemorrhage with or without associated edema is not considered a neurotoxicity feature and is excluded from ICANS grading. It may be graded according to CTCAE v5.0.  ASTCT: American Society for Transplantation and Cellular Therapy; CTCAE: Common Terminology Criteria for Adverse Events; EEG: electroencephalogram; ICANS: immune effector cell-associated neurological syndrome; ICE: immune-effector cell-associated encephalopathy; N/A: not applicable. | | | | |

**Supplementary Table 6. ICE Assessment Tool^6^**

| **Task** | **Details** | **Number of points** |
| --- | --- | --- |
| Orientation | Orientation to year, month, city, hospital | 4 points |
| Naming | Ability to name 3 objects (eg, point to clock, pen, button) | 3 points |
| Following commands | Ability to follow simple commands (eg, “Show me two fingers” or “Close your eyes and stick out your tongue”) | 1 point |
| Writing | Ability to write a standard sentence (eg, “Our national bird is the bald eagle”) | 1 point |
| Attention | Ability to count backwards from 100 by 10 | 1 point |
| Score interpretation: 10, no impairment; 7–9, grade 1 ICANS; 3–6, grade 2 ICANS; 0–2, grade 3 ICANS; 0, if patient is unarousable and unable to perform ICE assessment, grade 4 ICANS.  ICE: immune effector cell-associated encephalopathy; ICANS: immune effector cell-associated neurotoxicity syndrome. | | |

**Supplementary Table 7. Complete list of ICANS and associated neurological events^a^**

| 1. Acquired hepatocerebral degeneration  2. Alcoholic encephalopathy  3. Altered state of consciousness  4. Aphasia  5. Autoimmune encephalopathy  6. Brain oedema  7. Cerebellar cognitive affective syndrome  8. Chronic traumatic encephalopathy  9. Cognitive disorder  10. Cognitive linguistic deficit  11. Consciousness fluctuating  12. Contrast encephalopathy  13. Depressed level of consciousness  14. Diabetic encephalopathy  15. Encephalopathy  16. Encephalopathy allergic  17. Encephalopathy neonatal  18. Fumarase deficiency  19. Gerstmann Straussler Scheinker syndrome  20. Glutaric acidaemia type I  21. Hashimoto's encephalopathy  22. Hepatic encephalopathy  23. Hyperammonaemic crisis  24. Hyperammonaemic encephalopathy  25. Hyperbilirubinaemia neonatal  26. Hyperglycinaemia  27. Hypertensive encephalopathy  28. Hypoglycaemic encephalopathy  29. Hyponatraemic encephalopathy  30. Hypoxic-ischaemic encephalopathy neonatal  31. Hypoxic-ischaemic encephalopathy | 32. Idiopathic intracranial hypertension  33. Immune effector cell-associated neurotoxicity  Syndrome (ICANS)  34. Immune-mediated encephalopathy  35. Intracranial pressure increased  36. Kernicterus  37. Labrune syndrome  38. Leukoencephalopathy  39. Loss of consciousness  40. Marchiafava-Bignami disease  41. MELAS syndrome  42. Metabolic encephalopathy  43. Mitochondrial encephalomyopathy  44. Mitochondrial neurogastrointestinal encephalopathy  45. Muscular weakness  46. Opsoclonus myoclonus  47. Periventricular leukomalacia  48. Posterior reversible encephalopathy syndrome  49. Post-resuscitation encephalopathy  50. Purpura cerebri  51. Radiation-induced encephalopathy  52. Reye's syndrome  53. Seizure  54. Septic encephalopathy  55. Subacute myelo-opticoneuropathy  56. Toxic encephalopathy  57. Toxic leukoencephalopathy  58. Uraemic encephalopathy  59. Vascular cognitive impairment  60. Vasogenic cerebral oedema  61. Wernicke's encephalopathy |
| --- | --- |
| ^a^The list of ICANS and associated neurologic events were derived from a broad search using 61 selected preferred terms from MedDRA version 26.0.  ICANS, immune-effector cell-associated neurotoxicity syndrome; MedDRA, Medical Dictionary for Regulatory Activities. | |

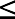
**Supplementary Table 8. Protocol recommendations for restarting tarlatamab after dosage delay^a^**

| **Last Dose**  **Administered** | **Time since last dose administered** | **Protocol recommended restarting guidelines^a^** |
| --- | --- | --- |
| **Step dose**  1 mg on  Cycle 1 Day 1 | ≤ 14 days | Proceed with target dose. |
|  | > 14 days | Repeat step dose schedule per cycle 1 guidelines, including pre-medication and hospitalization requirements. |
| **First target dose**  10 mg on  Cycle 1 Day 8 | ≤ 21 days | Proceed with target dose. |
|  | > 21 days | Repeat step dose schedule per cycle 1 guidelines, including pre-medication and hospitalization requirements. |
| **Any subsequent target dose** | ≤ 28 days | Proceed with target dose. |
|  | > 28 days | Repeat step dose schedule per cycle 1 guidelines, including pre-medication and hospitalization requirements. |

^a^If tarlatamab administration was delayed, the protocol defined restart guidelines for neurologic events, CRS, tumor lysis syndrome, non-febrile neutropenia, pituitary gland dysfunction, and hepatotoxicity. These restart guidelines were recommended for all other treatment interruptions and delays.

**Supplementary Table 9. Frequency of vital sign assessments in the first two cycles in the DeLLphi-301 trial**

| **Study Period** | **Frequency of Vital Sign Assessment** (Note: The frequency of vital sign assessment utilized in the DeLLphi-301 trial allowed for characterization of the CRS profile and may not reflect clinical practice) |
| --- | --- |
| **Cycle 1 (parts 1 and 2)** |  |
| All patients | - Every 15 (± 5) minutes during infusion and during first 2 hours post tarlatamab infusion - Every 30 (± 5) minutes from 2–4 hours post tarlatamab infusion - Every 1 hour (± 10) minutes from 4-8 hours post tarlatamab infusion - 12 hours (± 60 minutes) post tarlatamab infusion - 20 hours (± 60 minutes) post tarlatamab infusion - 24 hours (± 60 minutes) post tarlatamab infusion - After 24 hours post tarlatamab infusion, vital signs should be assessed per institutional standards |
| **Cycle 1 (Reduced monitoring substudy: Part 3)** | |
| All patients | - Every 30 (± 5) minutes during infusion and during first 4 hours post tarlatamab infusion - Every 1 hour (± 10) minutes from 4-8 hours post tarlatamab infusion - 12 hours (± 60 minutes) post tarlatamab infusion - 20 hours (± 60 minutes) post tarlatamab infusion - 24 hours (± 60 minutes) post tarlatamab infusion |
| **Cycle 2 (All Parts)** | |
| All patients | - Every 15 (± 5) minutes during infusion and during first 2 hours post tarlatamab infusion - Every 30 (± 5) minutes from 2–4 hours post tarlatamab infusion - Every 1 hour (± 10 minutes) from 4-8 hours post tarlatamab infusion |
| Patients that require hospitalization (additional time points) | - 12 hours (± 60 minutes) post tarlatamab infusion - 20 hours (± 60 minutes) post tarlatamab infusion - 24 hours (± 60 minutes) post tarlatamab infusion; after 24 hours post tarlatamab infusion, vital signs should be assessed per institutional standards |
| ^a^If a patient was not hospitalized on cycle 1 day 15, vital signs were only assessed up to 6 to 8 hours | |
